# Extending non-targeted exposure discovery of environmental chemical exposures during pregnancy and their association with pregnancy complications—a cross-sectional study

**DOI:** 10.1101/2022.03.07.22272040

**Authors:** Jessica Trowbridge, Dimitri Abrahamsson, Ting Jiang, Miaomiao Wang, June-Soo Park, Rachel Morello-Frosch, Marina Sirota, Dana E. Goin, Marya Zlatnick, Tracey J. Woodruff

**Affiliations:** Department of Obstetrics, Gynecology and Reproductive Sciences, Program on Reproductive Health and the Environment, University of California San Francisco, San Francisco 94143 California; Department of Toxic Substances Control, Environmental Chemistry Laboratory, California Environmental Protection Agency, Berkeley 94710, California; Department of Environmental Science and Policy Management and School of Public Health, University of California, Berkeley, Berkeley 94720 California; Bakar Computational Health Sciences Institute and Department of Pediatrics, University of California San Francisco, San Francisco 94158, California

**Keywords:** Non-targeted analysis, linear and branched PFOS, tridecanedioic acid, fatty acids, gestational diabetes mellitus, pregnancy hypertension

## Abstract

**Background:** Non-targeted Analysis (NTA) methods identify novel exposures; however, few chemicals have been quantified and interrogated with pregnancy complications.

**Objectives:** We characterize levels of nine exogenous and endogenous chemicals in maternal and cord blood identified, selected, and confirmed in prior NTA steps including: linear and branched isomers perfluorooctane sulfonate (PFOS); perfluorohexane sulfonate (PFHxS); monoethylhexyl phthalate; 4-nitrophenol; tetraethylene glycol; tridecanedioic acid, octadecanedioic acid; and deoxycholic acid. We evaluate relationships between maternal and cord levels and the relationship gestational diabetes mellitus (GDM) and hypertensive disorders of pregnancy in a diverse pregnancy cohort in San Francisco.

**Methods:** We collected matched maternal and cord serum samples from 302 pregnant people at delivery from the Chemicals in Our Bodies cohort in San Francisco. Chemicals were identified via NTA and quantified using targeted approaches. We calculate distributions and Spearman correlation coefficients testing the relationship of chemicals within and between the maternal and cord blood matrices. We used logistic regression to calculate the odds of GDM and hypertensive disorders of pregnancy associated with an interquartile range increase in maternal chemical exposures.

**Results:** We detected linear PFOS, PFHxS, octadecanedioic acid, and deoxycholic acid in at least 97% of maternal samples. Correlations ranged between -0.1 and 0.9. We observed strong correlations between cord and maternal levels of PFHxS (coefficient = 0.9), linear PFOS (0.8), and branched PFOS (0.8). An IQR increase in linear PFOS, branched PFOS, and octadecanedioic acid is associated with increased odds of GDM [OR (95%CI): 1.43 (0.96, 2.14), 1.56 (1.00, 2.44), and 1.26 (0.83, 1.92) respectively] and tridecanedioic acid positively associated with hypertensive disorders of pregnancy [1.28 (0.90, 1.86)].

**Discussion:** We identified both exogenous and endogenous chemicals, two of which (octadecanedioic acid and tridecanedioic acid) have both endogenous and exogenous sources, and which have seldom been quantified in pregnant people or related to pregnancy complications.

## 1. Introduction

Prenatal exposures to environmental chemicals are ubiquitous in the United States (U.S.)^1^ and studies have demonstrated they can have lifelong consequences for maternal and child health outcomes,^2^ such as neurodevelopment, cardiovascular disease, and reproduction. However, fewer than 1% of over 80,000 chemicals produced and used in the U.S. are regularly biomonitored^3^ and fewer still have been assessed for adverse health outcomes during pregnancy. Non-targeted analysis (NTA) coupled with a prioritization step and quantification using targeted methods can facilitate the identification and quantitation of previously unmeasured chemical exposures.^4^ Additionally, by identifying and measuring novel chemical exposures we can investigate their relationship with pregnancy complications, facilitating the identification of opportunities for prevention.

Increasingly, studies are applying NTA to better characterize exposure to toxic environmental chemicals seldom measured in pregnant people.^5,6^ NTA studies in pregnant and non-pregnant populations have identified both known and novel chemical exposures, including PFAS congeners, pesticides, flame retardants, and other industrial chemicals, which have been linked with pregnancy complications.^6,7^ For example, PFAS has been linked with increased risk of miscarriage and preeclampsia,^8^ and chemicals with fatty acid-like structures, including PFAS and other long-chain hydrocarbons, have been shown to interact with endogenous fatty acid metabolism impacting placental development and may contribute to pregnancy complications and adverse birth outcomes.^6,9–11^ Similarly, NTA studies have been able to link novel chemical exposures and metabolism biomarkers.^6,12^ However, the vast majority of novel chemicals identified through NTA have not been evaluated for associations with pregnancy related complications.

This study builds on previous NTA work done on the Chemicals in Our Bodies-2 (CIOB2) cohort, in which we first identified chemical features using high resolution mass spectrometry (HRMS) and confirmed chemical identities via comparison to a standard.^5,6,13^ The aim of the present study is to describe the quantification of a subset of chemicals identified and prioritized through this prior NTA and to evaluate relationships between chemical levels and pregnancy complications. Supplementing the non-targeted discovery of chemicals with targeted quantification described in this analysis addresses some of the limitations of NTA; namely, targeted approaches increase the sensitivity to measure and compare chemical concentrations and facilitate assessment of relationships between chemicals and pregnancy complications.

## 2. Methods

### 2.1 Data and Sample Collection

We enrolled pregnant participants in the CIOB2 cohort between 2014 and 2018 from the prenatal clinics at the Zuckerberg San Francisco General Hospital (SFGH) and Mission Bay or Moffit Long (MB/ML) hospitals. Participant recruitment and sample collection is described elsewhere.^4,5,14^ Briefly, participants were recruited during the second trimester of pregnancy, with eligibility criteria including 18-40 years of age, English or Spanish speaking, with singleton pregnancies, and no diagnosed pregnancy complications at recruitment. Paired maternal and umbilical cord blood were collected from participants who agreed to bank their samples and be contacted for participation in future studies. Maternal samples were collected at delivery and cord blood was collected after delivery but before clamping whenever possible. Blood was collected in BD Vacutainer Plus serum tubes, spun at 3000 rpm to collect the serum, and stored at -80°C until analysis. Study protocols were approved by the Institutional Review Boards at the University of California, San Francisco and Berkeley (# 13-12160).

Demographic information was collected via interviews conducted during the second trimester of pregnancy and included maternal age, race and ethnicity (white, Latinx, and Non-Hispanic Asian/Pacific Islander, other race—due to low numbers of Black, Native American, and multiracial participants, these groups were collapsed into “other” for model specification), maternal education (some college or less, completed a college degree, >college degree), household income (<40k per year, 40 to 100k, >100k), nativity (born in the U.S. yes or no). Pre-pregnancy body mass index (BMI kg/m2; <18.5, 18.5 to <25, 25 to <35, >=35)^15^ and hospital of delivery [Zuckerberg San Francisco General Hospital (SFGH) or Mission Bay/ Moffit Long (MB/ML)] were abstracted from the medical record.

Pregnancy complications were identified by clinical diagnoses and abstracted from the medical record. These included the binary outcomes of: gestational diabetes mellitus (GDM), preeclampsia, and gestational hypertension. Preeclampsia and pregnancy related hypertension were combined into one outcome called hypertensive disorder of pregnancy because they make up part of a larger spectrum of pregnancy complications related to hypertension.^16,17^

### 2.2 Chemical selection

Nine environmental chemicals were measured in maternal and cord samples using targeted approaches described below. Linear and branched isomers of perfluorooctane sulfonate (PFOS); perfluorohexane sulfonate (PFHxS); monoethylhexyl phthalate (MEHP), a metabolite of diethylhexyl phthalate, a common plasticizer^18^; 4-nitrophenol, a known endocrine disruptor used in pesticides, dyes, and pharmaceuticals^19^; tetraethylene glycol, a plasticizer and solvent used in metal lubricants, printing inks, and cosmetics;^20^ tridecanedioic acid and octadecanedioic acid which are endogenous fatty acids that also have exogenous sources primarily from plastics synthesis;^21–23^ and deoxycholic acid, a bile acid and endogenous biomarker associated with GDM **(Table 1)**.^24^

**Table 1.**
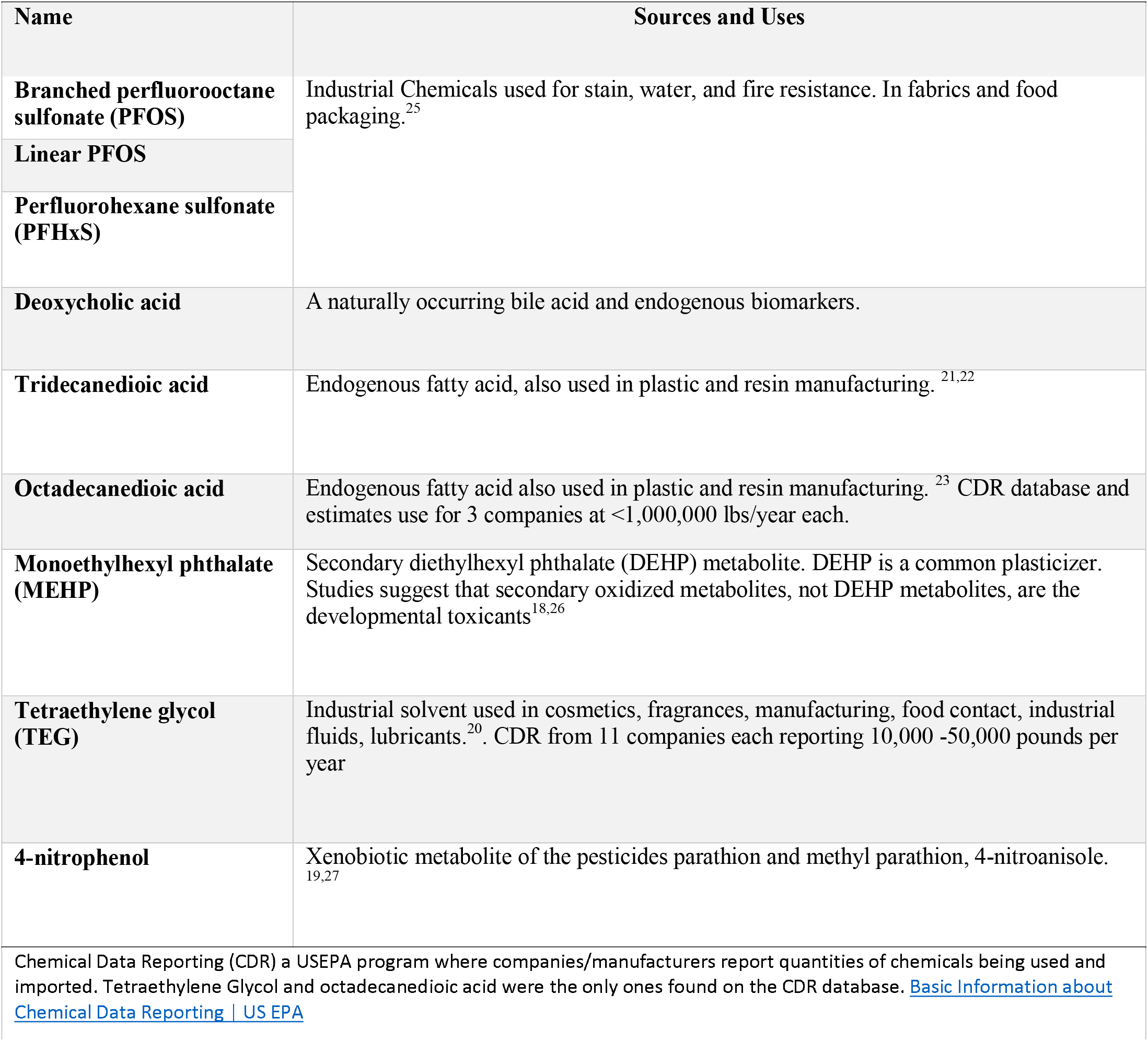
Chemical uses and sources of the nine chemicals identified from non-targeted analyses (NTA) and selected for confirmation and quantification

| Name | Sources and Uses |
| --- | --- |
| **Branched perfluorooctane sulfonate (PFOS)** | Industrial Chemicals used for stain, water, and fire resistance. In fabrics and food packaging.<sup>25</sup> |
| **Linear PFOS** |  |
| **Perfluorohexane sulfonate (PFHxS)** |  |
| **Deoxycholic acid** | A naturally occurring bile acid and endogenous biomarkers. |
| **Tridecanedioic acid** | Endogenous fatty acid, also used in plastic and resin manufacturing.<sup>21,22</sup> |
| **Octadecanedioic acid** | Endogenous fatty acid also used in plastic and resin manufacturing.<sup>23</sup> CDR database and estimates use for 3 companies at <1,000,000 lbs/year each. |
| **Monoethylhexyl phthalate (MEHP)** | Secondary diethylhexyl phthalate (DEHP) metabolite. DEHP is a common plasticizer. Studies suggest that secondary oxidized metabolites, not DEHP metabolites, are the developmental toxicants<sup>18,26</sup> |
| **Tetraethylene glycol (TEG)** | Industrial solvent used in cosmetics, fragrances, manufacturing, food contact, industrial fluids, lubricants.<sup>20</sup>. CDR from 11 companies each reporting 10,000 -50,000 pounds per year |
| **4-nitrophenol** | Xenobiotic metabolite of the pesticides parathion and methyl parathion, 4-nitroanisole.<sup>19,27</sup> |

Chemical Data Reporting (CDR) a USEPA program where companies/manufacturers report quantities of chemicals being used and imported. Tetraethylene Glycol and octadecanedioic acid were the only ones found on the CDR database. Basic Information about Chemical Data Reporting | US EPA

The selection and confirmation procedures for selection of these chemicals is described elsewhere.^5,6,13^ Briefly, in previous NTA studies, 30 matched maternal and cord samples were scanned using HRMS in positive and negative ionization modes identifying suspect features. Detected features were matched to an in-house database and features were then prioritized for MS/MS fragmentation based on the ubiquity of exposure, feature intensity differences across demographic variables, and correlation between maternal and cord samples. Fifteen chemicals were confirmed at level 1 confidence via comparison to analytical standards.^28^ Level 1 confirmed features included monoethylhexyl phthalate, 4-nitrophenol, tridecanedioic acid, and octadecanedioic acid. Subsequent generalized suspect screening analysis of this cohort was expanded to include 295 matched maternal and cord samples and a more comprehensive database including both endogenous and exogenous compounds. This resulted in nineteen chemicals confirmed with analytical standards to level 1 confidence, including those previously identified, and adding PFHxS, PFOS, and deoxycholic acid which were tentatively identified through MS/MS fragmentation (confidence level 2). We selected chemicals for targeted quantification if the chemical was identified in previous NTA with level 1 or level 2 confidence, is a widely used industrial compound with high potential for exposure, is associated with pregnancy complications, or is not regularly biomonitored by the Center for Disease Control or other large biomonitoring studies.^5^

### 2.3 Targeted chemical analysis

Quantitation analysis of the nine selected chemicals was carried out using Agilent MassHunter Quantitative Analysis software package (Agilent Technologies Inc., Version 10.0). Analytical grade standards were purchased from suppliers (Sigma Aldrich and Thermo Fisher) and serial dilutions of the standards were prepared in methanol. Calibration curve of each of the specific standard was acquired under the same condition as serum sample analysis, using the LC/QTOF-MS (Agilent 1290 UPLC interfaced with Agilent iFunnel 6550 QTOF-MS system) as described previously.^5,6,13^ M2-PFOA (Wellington Laboratories, Ontario, Canada) was used as internal standard for negative ionization chemicals, while D15-TPP (Cambridge Isotope Laboratories, Tewksbury, MA) for positive ionization chemicals. Particularly, PFOS in serum samples were separated chromatically, thus both total and linear PFOS quantitation were calculated, and branched PFOS concentrations were obtained by subtracting linear portion from the total PFOS.

### 2.4 Statistical analysis

To address chemical levels below the limit of detection we used machine reported values when available or, if not available, we imputed levels below the LOD with LOD/sqrt(2). We assessed the detection frequency (percent above the limit of detection) and limited our statistical analysis to chemicals with a detection frequency of 67% or higher. We described the distribution of chemicals in maternal and cord blood samples, and calculated Spearman correlation coefficients for chemicals within and between maternal and cord samples. We natural log-transformed the data for use in statistical models and applied logistic regression models to assess the relationship between maternal chemical exposure and pregnancy complications. Potential confounders were identified *a priori* (**Supplemental Figure 1**), and our final models were adjusted for maternal age, hospital of delivery, and race/ethnicity. We considered hospital of delivery as a cumulative indicator of socioeconomic status (including income, educational attainment, and insurance status). Patients in both hospitals are racially and ethnically diverse. However, SFGH is a safety net hospital that serves a large proportion of patients without private health insurance, while MB/ML serves patients of higher socioeconomic status and with private health insurance. Race and ethnicity are included in models as an indicator of the exposure to structural racism, that is independently associated with chemical exposure and adverse pregnancy complications. We calculated the interquartile range (IQR) of each natural log transformed chemical. Beta coefficients and 95% confidence intervals (CI) were multiplied IQR and exponentiated to identify the odds ratio (OR) of each pregnancy complication for each IQR increase in exposure level.

## 3. Results

### Participants, demographics, and medical record data

The analysis included 302 participants (**Table 2**). The average age at delivery was 33 years of age. Forty-three percent of participants identified as white and 32% as Latinx or Hispanic. Most participants completed college or graduate degrees (>65%), were born in the U.S. (54%), and had a household income of over $100,000 per year (56%). For the outcomes, 21% of participants had a diagnosis of GDM, 13% gestational hypertension, and 7% preeclampsia.

**Table 2.** Demographic variables and outcomes in Chemicals in Our Bodies 2 participants (n= 302) recruited between 2014 and 2018 in San Francisco

|  | Mean(SD) or N(%) |
| --- | --- |
| **COVARIATES** |  |
| **Maternal age at delivery (years)** | 33 (±5) |
| **Race/ethnicity** |  |
| White | 130 (43%) |
| Latinx | 96 (32%) |
| Asian | 47 (16%) |
| Other or Unknown <sup>a</sup> | 29 (10%) |
| **Education** |  |
| Some college or less | 94 (31%) |
| College degree | 73 (24%) |
| Graduate degree | 125 (41%) |
| Missing | 10 (3%) |
| **Household income** |  |
| <40k | 62 (21%) |
| 40k-100k | 36 (12%) |
| >100k | 170 (56%) |
| Missing | 34 (11%) |
| **Born in U.S.** |  |
| No | 110 (36%) |
| Yes | 147 (49%) |
| Missing | 45 (15%) |
| **Pre-pregnancy BMI <sup>b</sup>** |  |
| <18.5 | 6 (2%) |
| 18.5 to <25 | 147 (49%) |
| 25 to <30 | 73 (24%) |
| 30 or higher | 41 (14%) |
| Missing | 35 (12%) |
| **Hospital of Delivery** |  |
| San Francisco General Hospital | 88 (29%) |
| Mission Bay | 213 (71%) |
| Missing | 1 (<1%) |
| **OUTCOMES** |  |
| **Gestational diabetes mellitus** |  |
| No | 237 (78%) |
| Yes | 63 (21%) |
| Missing | 2 (1%) |
| **Preeclampsia** |  |
| No | 268 (89%) |
| Yes | 20 (7%) |
| Missing | 14 (5%) |
| **Gestational hypertension** |  |
| No | 248 (82%) |
| Yes | 39 (13%) |
| Missing | 15 (5%) |
| **Hypertensive disorders of pregnancy <sup>c</sup>** |  |
| None | 238 (79%) |
| Any | 50 (17%) |
| Missing | 14 (5%) |

<sup>a</sup>Race and ethnicity categories were collapsed for use in linear models due to small numbers in some categories. Other/unknown includes Black, Native Hawaiian or Pacific Islander, Native American, multiracial and individuals of unknown race; <sup>b</sup> BMI is calculated as kg/m<sup>2</sup> and cut-offs are based on the CDC guidelines;<sup>15</sup>
<sup>c</sup> Combines preeclampsia and gestational hypertension.

### 3.1 Distribution and correlation of chemical levels in maternal samples

Participants had at least four chemicals in maternal samples and three chemicals in cord samples. PFHxS, linear PFOS, deoxycholic acid, and octadecanedioic acid were all detected in at least 97% of maternal samples. Deoxycholic acid, tridecanedioic acid, PFHxS, were detected in at least 87% of cord blood samples (Table 3). Since we estimated branched PFOS levels by subtracting linear PFOS from the total PFOS we did not calculate a detection frequency for this chemical. Chemical detection frequency varied between maternal and cord blood samples. Linear PFOS, and 4-nitrophenol were detected more often maternal samples while monoethylhexyl phthalate, and tridecanedioic acid were detected more frequently in cord blood. Similarly, the average chemical levels varied across the different matrices with branched PFOS, linear PFOS, deoxycholic acid, and octadecanedioic acid found at higher levels in maternal samples compared to cord samples, while tridecanedioic acid was higher in cord than in maternal samples.

**Table 3.**
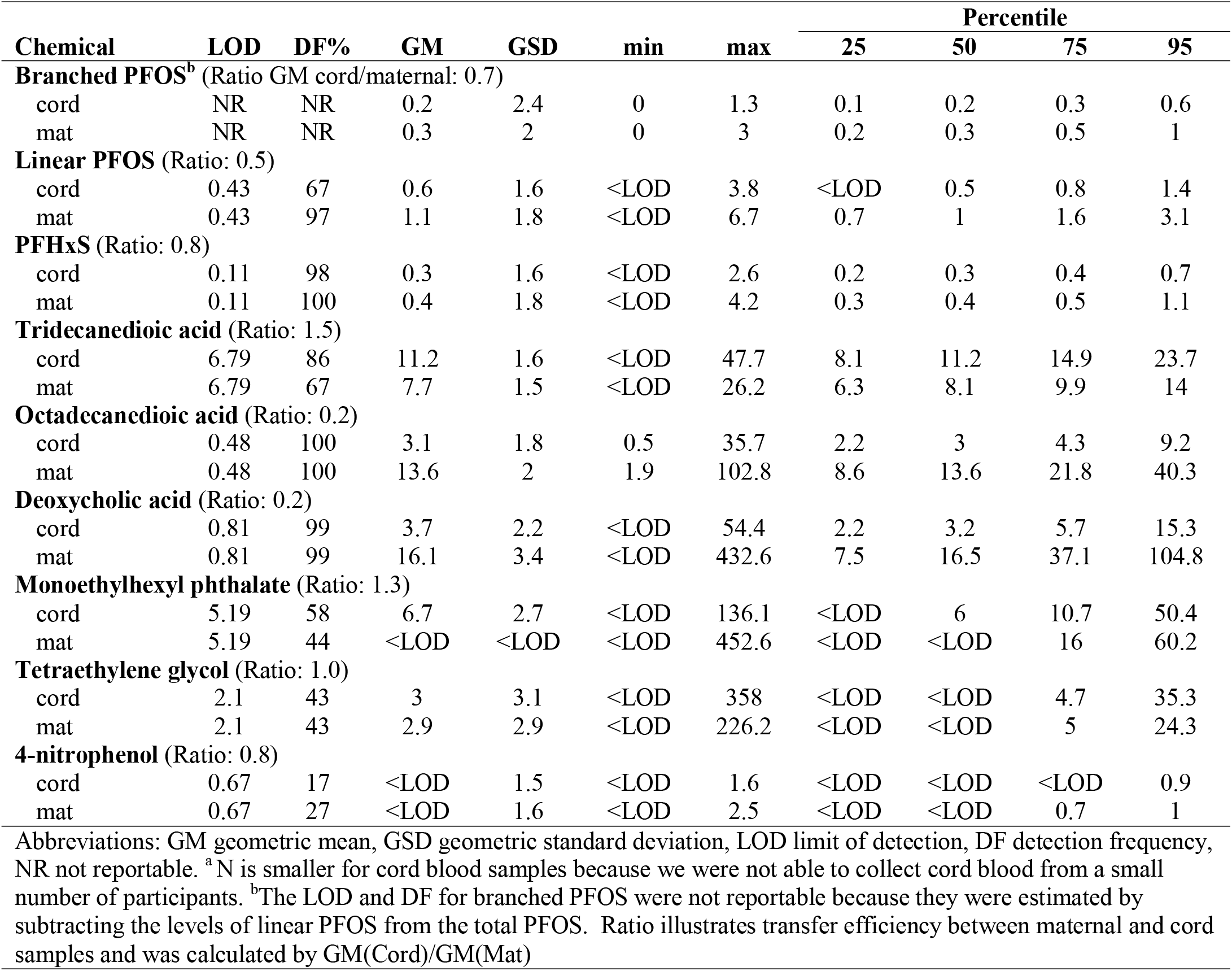
Distribution of chemical levels in maternal serum (n= 302) and cord blood^a^ (n= 299) (ng/ml)

| Chemical | LOD | DF% | GM | GSD | min | max | Percentile |  |  |  |
| --- | --- | --- | --- | --- | --- | --- | --- | --- | --- | --- |
|  |  |  |  |  |  |  | 25 | 50 | 75 | 95 |
| **Branched PFOS<sup>b</sup>** (Ratio GM cord/maternal: 0.7) |  |  |  |  |  |  |  |  |  |  |
| cord | NR | NR | 0.2 | 2.4 | 0 | 1.3 | 0.1 | 0.2 | 0.3 | 0.6 |
| mat | NR | NR | 0.3 | 2 | 0 | 3 | 0.2 | 0.3 | 0.5 | 1 |
| **Linear PFOS** (Ratio: 0.5) |  |  |  |  |  |  |  |  |  |  |
| cord | 0.43 | 67 | 0.6 | 1.6 | <LOD | 3.8 | <LOD | 0.5 | 0.8 | 1.4 |
| mat | 0.43 | 97 | 1.1 | 1.8 | <LOD | 6.7 | 0.7 | 1 | 1.6 | 3.1 |
| **PFHxS** (Ratio: 0.8) |  |  |  |  |  |  |  |  |  |  |
| cord | 0.11 | 98 | 0.3 | 1.6 | <LOD | 2.6 | 0.2 | 0.3 | 0.4 | 0.7 |
| mat | 0.11 | 100 | 0.4 | 1.8 | <LOD | 4.2 | 0.3 | 0.4 | 0.5 | 1.1 |
| **Tridecanedioic acid** (Ratio: 1.5) |  |  |  |  |  |  |  |  |  |  |
| cord | 6.79 | 86 | 11.2 | 1.6 | <LOD | 47.7 | 8.1 | 11.2 | 14.9 | 23.7 |
| mat | 6.79 | 67 | 7.7 | 1.5 | <LOD | 26.2 | 6.3 | 8.1 | 9.9 | 14 |
| **Octadecanedioic acid** (Ratio: 0.2) |  |  |  |  |  |  |  |  |  |  |
| cord | 0.48 | 100 | 3.1 | 1.8 | 0.5 | 35.7 | 2.2 | 3 | 4.3 | 9.2 |
| mat | 0.48 | 100 | 13.6 | 2 | 1.9 | 102.8 | 8.6 | 13.6 | 21.8 | 40.3 |
| **Deoxycholic acid** (Ratio: 0.2) |  |  |  |  |  |  |  |  |  |  |
| cord | 0.81 | 99 | 3.7 | 2.2 | <LOD | 54.4 | 2.2 | 3.2 | 5.7 | 15.3 |
| mat | 0.81 | 99 | 16.1 | 3.4 | <LOD | 432.6 | 7.5 | 16.5 | 37.1 | 104.8 |
| **Monoethylhexyl phthalate** (Ratio: 1.3) |  |  |  |  |  |  |  |  |  |  |
| cord | 5.19 | 58 | 6.7 | 2.7 | <LOD | 136.1 | <LOD | 6 | 10.7 | 50.4 |
| mat | 5.19 | 44 | <LOD | <LOD | <LOD | 452.6 | <LOD | <LOD | 16 | 60.2 |
| **Tetraethylene glycol** (Ratio: 1.0) |  |  |  |  |  |  |  |  |  |  |
| cord | 2.1 | 43 | 3 | 3.1 | <LOD | 358 | <LOD | <LOD | 4.7 | 35.3 |
| mat | 2.1 | 43 | 2.9 | 2.9 | <LOD | 226.2 | <LOD | <LOD | 5 | 24.3 |
| **4-nitrophenol** (Ratio: 0.8) |  |  |  |  |  |  |  |  |  |  |
| cord | 0.67 | 17 | <LOD | 1.5 | <LOD | 1.6 | <LOD | <LOD | <LOD | 0.9 |
| mat | 0.67 | 27 | <LOD | 1.6 | <LOD | 2.5 | <LOD | <LOD | 0.7 | 1 |

Abbreviations: GM geometric mean, GSD geometric standard deviation, LOD limit of detection, DF detection frequency, NR not reportable. <sup>a</sup>N is smaller for cord blood samples because we were not able to collect cord blood from a small number of participants. <sup>b</sup>The LOD and DF for branched PFOS were not reportable because they were estimated by subtracting the levels of linear PFOS from the total PFOS. Ratio illustrates transfer efficiency between maternal and cord samples and was calculated by GM(Cord)/GM(Mat)

Monoethylhexyl phthalate, tetraethylene glycol, and 4-nitrophenol were each detected in fewer than 50% of maternal samples, however 4-nitrophenol was detected in more maternal serum samples than cord samples, while monoethylhexyl phthalate was found in more cord blood samples. Due to their low detection frequency, monoethylhexyl phthalate, tetraethylene glycol, and 4-nitrophenol, were excluded from further analyses.

When assessing the Spearman correlation coefficients (**Figure 2**) of the same chemical across maternal and cord samples, we found that linear PFOS, and branched PFOS and PFHxS were highly correlated in the maternal and cord samples (correlation coefficients of 0.8, 0.8, and 0.9 respectively). When looking at the correlations of different chemicals in the maternal and cord samples we found moderate correlations between cord-linear PFOS and maternal-branched PFOS, as well as between cord-linear PFOS and maternal-PFHxS (coefficients 0.7 and 0.5 respectively). In the cord blood samples, linear PFOS was highly correlated with branched PFOS and moderately with PFHxS (coefficients of 0.8 to 0.6 respectively), and octadecanedioic acid was moderately correlated with tridecanedioic acid (coefficient of 0.4). In the maternal samples, the relationship between linear and branched PFOS had a correlation coefficient of 0.8, branched PFOS and PFHxS a correlation of 0.6, and between linear PFOS and PFHxS of 0.6. The PFAS chemicals were weakly correlated with the fatty acids—octadecanedioic acid, tridecanedioic acid—and neither PFAS, nor the fatty acids were correlated with deoxycholic acid, within or between the maternal serum and cord blood samples.

**Figure 1.**
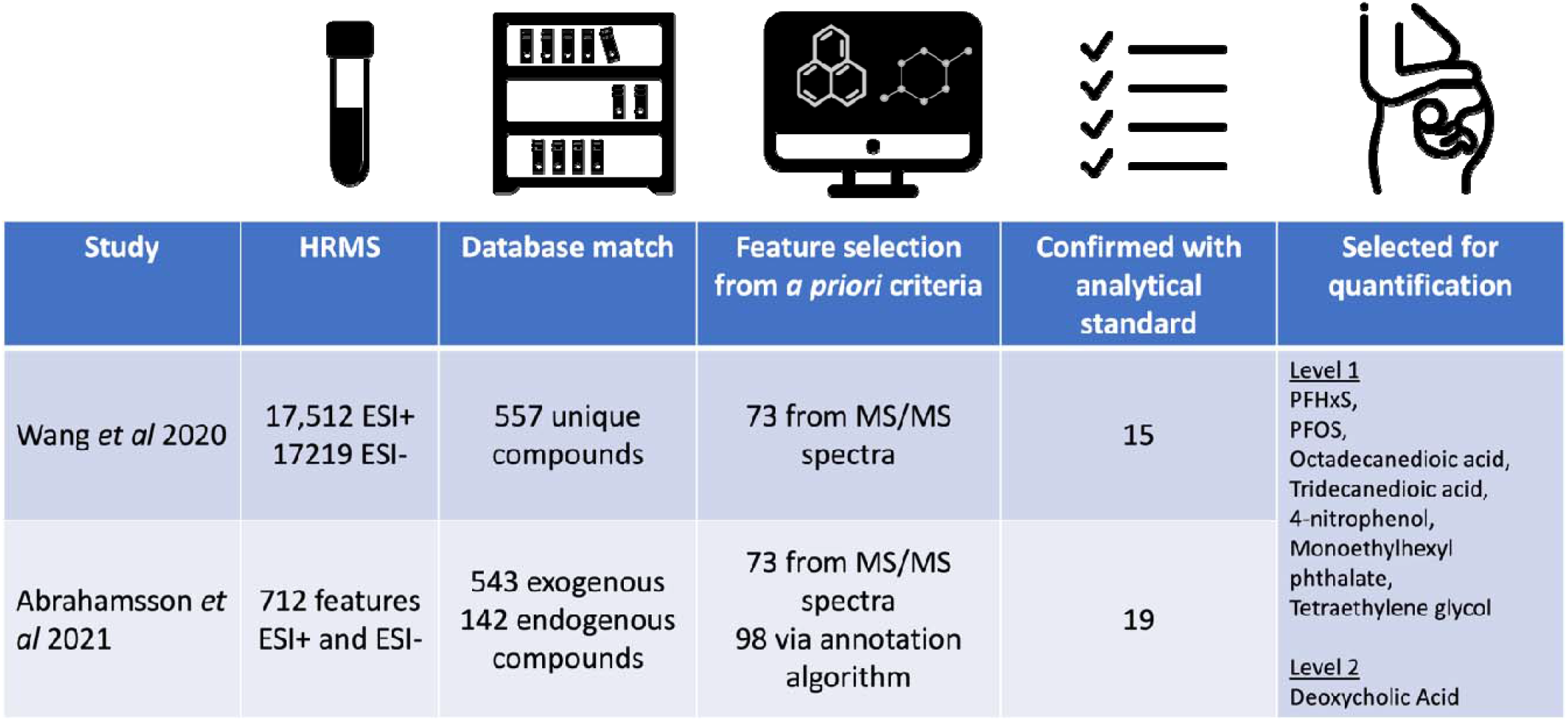
Summary of NTA procedures leading to the selection of nine chemical for quantification.

**Figure 2:**
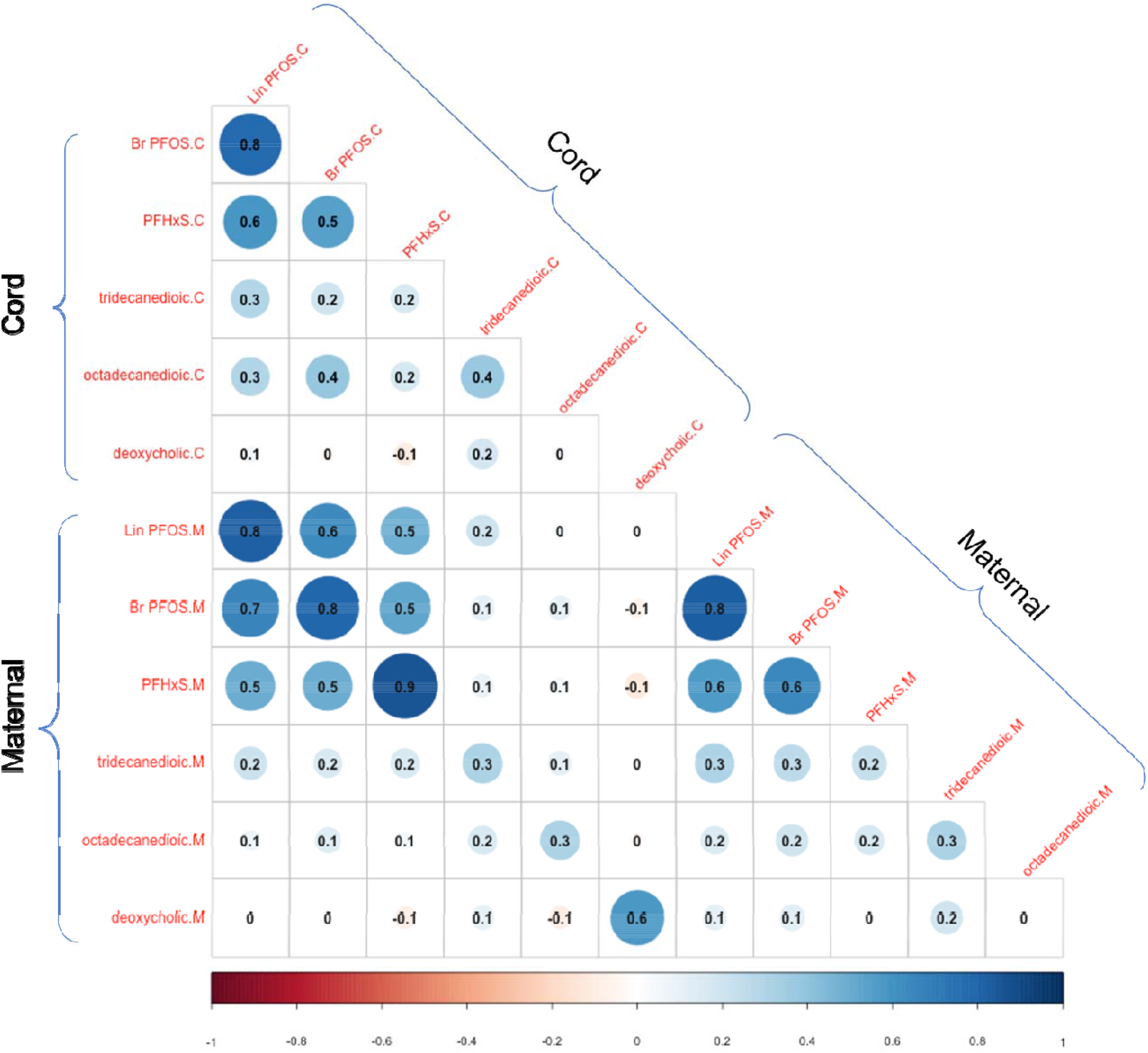
Spearman correlation coefficients for chemicals detected in maternal and cord samples, chemical have at least 65% DF in maternal and cord samples (n = 302).

### 3.2 Association between chemical exposure with pregnancy complications

From our logistic models we observed a trend of increased odds of GDM for each IQR increase in levels of exposure to environmental chemicals and increased odds of pregnancy hypertensive disorders with tridecanedioic acid (Table 4). For the relationship between chemical exposures and GDM, all the chemicals included in this analysis had an OR above 1. Stronger associations were observed for branched PFOS, linear PFOS, and octadecanedioic acid; The unadjusted relationship between linear PFOS and branched PFOS with GDM was statistically significant. The direction and measures of association we observed between chemical levels and hypertensive disorders of pregnancy was less consistent than for GDM. Exposure to tridecanedioic acid was associated with an increase in the odds of hypertensive disorders of pregnancy [OR (95%CI): 1.28(0.90, 1.86)]. Exposure to tridecanedioic acid was also associated in increased odds of the individual outcomes that made up hypertensive disorders of pregnancy—preeclampsia and gestational hypertension (**Supplemental Table 1**). The ORs for PFHxS, octadecanedioic acid, and deoxycholic acid with gestational hypertension approximated 1. Linear and branched PFOS had an OR of <1 but the confidence intervals included the null value.

**Table 4.** Adjusted and unadjusted odds ratios and 95% confidence intervals from linear models of the relationship between an interquartile range (IQR) chemical exposure at delivery and pregnancy complications (n = 302)

| Chemical | OR (95% CI) |  |
| --- | --- | --- |
|  | gestational diabetes | hypertensive disorders <sup>b</sup> |
| Number missing | 2 | 14 |
| N(%) with outcome | 63 (21%) | 50 (17%) |
| **Branched PFOS** |  |  |
| Model0 | 1.65 ( 1.10 , 2.52 ) | 1.05 ( 0.68 , 1.62 ) |
| Model1 | 1.56 ( 1.00 , 2.46 ) | 1.06 ( 0.65 , 1.71 ) |
| **Linear PFOS** |  |  |
| Model0 | 1.50 ( 1.05 , 2.17 ) | 0.85 ( 0.56 , 1.27 ) |
| Model1 | 1.43 ( 0.96 , 2.14 ) | 0.89 ( 0.56 , 1.39 ) |
| **PFHxS** |  |  |
| Model0 | 1.15 ( 0.86 , 1.51 ) | 1.00 ( 0.72 , 1.35 ) |
| Model1 | 1.09 ( 0.80 , 1.48 ) | 0.98 ( 0.67 , 1.38 ) |
| **Octadecanedioic acid** |  |  |
| Model0 | 1.20 ( 0.82 , 1.77 ) | 1.05 ( 0.69 , 1.63 ) |
| Model1 | 1.26 ( 0.83 , 1.92 ) | 1.00 ( 0.65 , 1.54 ) |
| **Tridecanedioic acid** |  |  |
| Model0 | 1.08 ( 0.79 , 1.49 ) | 1.25 ( 0.88 , 1.80 ) |
| Model1 | 1.03 ( 0.74 , 1.45 ) | 1.28 ( 0.90 , 1.86 ) |
| **Deoxycholic acid** |  |  |
| Model0 | 1.16 ( 0.81 , 1.69 ) | 0.96 ( 0.65 , 1.43 ) |
| Model1 | 1.17 ( 0.80 , 1.72 ) | 0.97 ( 0.65 , 1.45 ) |

<sup>a</sup> CI = Confidence Interval; Model0 is the unadjusted model; Model1 is adjusted for maternal age, hospital of delivery, and race/ethnicity; <sup>b</sup> includes both preeclampsia and gestational hypertension

## 4. Discussion

Building on NTA steps from prior studies^4,6,13^, we confirmed and quantified the presence of nine chemicals, some of which have not been previously measured in pregnant people, such as tridecanedioic acid, octadecanedioic acid, and tetraethylene glycol, or rarely measured in pregnant people like 4-nitrophenol. In addition, a subset of these were moderately to strongly associated with GDM and hypertensive disorders of pregnancy. Linear and Branched PFOS were associated with increased odds of GDM and tridecanedioic acid was associated with hypertensive disorders of pregnancy. This study adds to our understanding of the associations between environmental exposures during pregnancy and pregnancy complications.

This study provides further evidence of prenatal exposure to environmental chemicals, some of which have positive associations with pregnancy complications. A 2020 study suggests that PFAS exposure during pregnancy may also affect metabolic pathways that adversely impact placental development^9^ and while evidence remains mixed, PFAS has been shown to be associated with hypertensive disorders of pregnancy, preeclampsia, and GDM.^9,29–31^ The linear and branched forms of PFOS have not previously been well characterized during pregnancy. Furthermore, metabolism, excretion, and placental transfer may vary by the linear and branched forms of this ubiquitous chemical and lead to different health outcomes and pregnancy complications.^25^

Significant positive correlations between branched PFOS, linear PFOS and PFHxS in the maternal and cord samples suggest that they may share a common source of exposure. Indeed, traditional PFOS production yielded a mixture of branched and linear isomers with approximately 30% of isomers in the branched form.^25^ From linear models we found that the linear and branched forms of PFOS may have a relationship with pregnancy complications. Branched and Linear PFOS were significantly associated with GDM in unadjusted models and although the relationship was attenuated in adjusted models, the direction and magnitude of the association were maintained. Scientific evidence indicates transplacental transfer of PFAS chemicals^6,32,33^ and gestational diabetes may facilitate transplacental transfer of PFOS.^34^ Linear PFOS makes up a larger proportion of the measured PFOS,^25^ is hypothesized to remain longer in maternal serum and is therefore more available in maternal samples than branched PFOS, which may contribute to the differences in association we observed with GDM.^35,36^

Previous studies have found that increased exposure to fatty acids may be associated with an increased risk of hypertensive disorders of pregnancy due to disruption of fatty acid regulation and metabolism.^37^ We saw an association between tridecanedioic acid and hypertensive disorders of pregnancy, but not for octadecanedioic acid. Similarly, although the bile acid, deoxycholic acid, is associated GDM in other studies,^24^ we did not see elevated risk of these pregnancy complications associated with deoxycholic acid in our study. The endogenous chemicals in our study have seldom been studied in pregnant women and the fatty acids are also known to have exogenous sources and uses.

Tetraethylene glycol, 4-nitrophenol and monoethylhexyl phthalate were present in a small subset of our participants but were not evaluated for an association with pregnancy complications because their detection frequency in maternal samples was below 65%. In addition, these chemicals have short half-lives and are quickly metabolized and excreted in urine contributing to their low detection frequency in serum samples.^19,26,38^ These are high production volume chemicals that may interact with endocrine and metabolic pathways; however, their relationship with adverse reproductive or pregnancy outcomes has not been previously assessed, warranting further study. Despite their low detection frequency and that they were measured in serum rather than in urine our study demonstrates that novel exposures to these environmental chemicals can still be identified through non-targeted methods and confirmed through targeted approaches.

There are several limitations that may influence the associations we observed in our study. First is the timing of the collection of samples and assessment of health outcomes. This is a cross-sectional study with samples collected at delivery and pregnancy complications were identified from clinical diagnosis abstracted from medical record. Because of this we cannot assess temporality of our exposures and outcome. Nevertheless, the chemicals included in this study are industrial chemicals, many of which are high production volume chemicals that have a high potential for exposure. For example, PFAS chemicals are ubiquitous in biological matrices and the environment, and tridecanedioic acid and octadecanedioic acid are used in the synthesis of plastics and are considered high production volume chemicals.^21^ While the chemical measurements reflect a snapshot of an individual’s exposure history during pregnancy the sources and uses of these environmental chemicals imply that there is a potential for prolonged exposure throughout the pregnancy. Studies show that PFAS levels decrease in later trimesters of pregnancy,^39,40^ thus the PFAS levels we measured at delivery may be lower than in other trimesters of pregnancy biasing our results toward to the null. Another potential limitation is that levels of branched PFOS were estimated and not quantified with a standard and thus the levels of branched PFOS may reduce precision of calculated measures of association. Likewise, the low detection frequency of 4-nitrophenol, tetraethylene glycol, or monoethylhexyl phthalate precluded our ability to assess the chemical-outcome relationship. However, these chemicals remain understudied in pregnant people and are of high concern due to their widespread use as industrial chemicals and limited understanding of their health effects. Finally, because our recruitment criteria indicated that participants with pregnancy complications by the second trimester we may have missed earlier manifestations of pregnancy complications. This could undercount the number of GDM and hypertensive disorders of pregnancy biasing our results toward the null. However, diagnosis of hypertensive disorders of pregnancy and testing for GDM usually occurs in the third trimester of pregnancy while our participants were recruited in the second.^41,42^

In this analysis we identified and quantified exogenous and endogenous environmental chemicals, some of which have not been previously measured during pregnancy nor evaluated for associations with pregnancy complications. Our study supports prior reports of associations between the branched and linear forms of PFOS and GDM and adds to the limited research on exposure to tridecanedioic acid and octadecanedioic acid, endogenous fatty acids that also have industrial sources. Finally, this study further illustrates the benefit of pairing non-targeted and targeted approaches to identify, confirm, and quantify novel chemical exposures. Combining NTA methods with targeted chemicals measurement can be further applied to identifying, quantifying, and evaluating metabolism biomarkers that are on the pathway between exposure and downstream adverse health effects.

## Data Availability

Data is not available due to IRB limitatations

