## Supplemental table and figure for "Extending non-targeted exposure discovery of environmental chemical exposures during pregnancy and their association with pregnancy complications—a cross-sectional study"

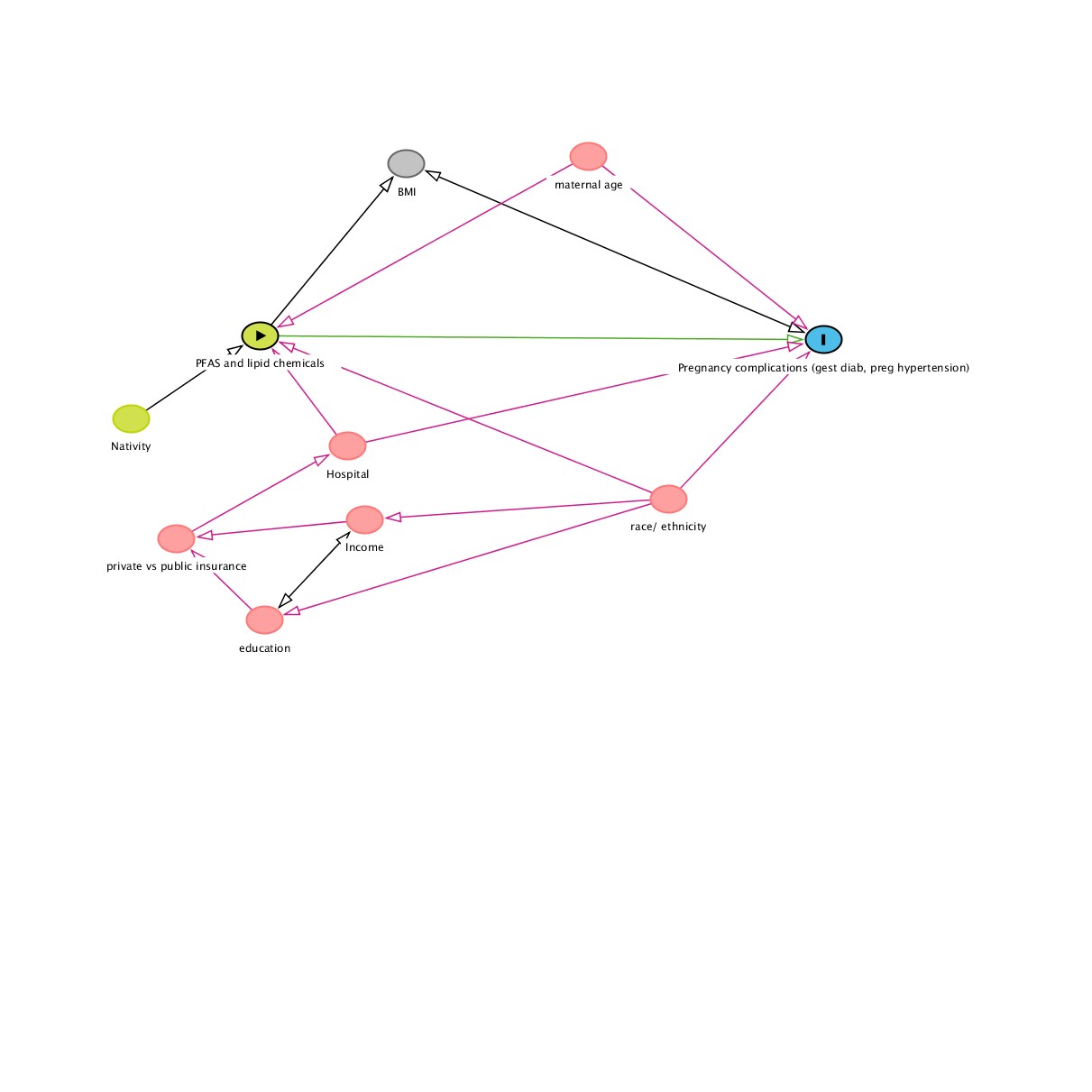

Figure S1. Directed Acyclic Graph (DAG) of the relationship between chemical exposures and pregnancy complications, identifying confounders of interest.

| S. Table 2. Odds Ratios from unadjusted and adjusted models for the relationship between chemical exposure during the 2nd trimester and preeclampsia and gestational hypertension | | | |
| --- | --- | --- | --- |
|  |  | **OR (95% CI)** | |
| **Chemical** |  | **Preeclampsia** | **Gestational Hypertension** |
|  | | missing n= 14 | missing n= 15 |
| **N(%)** |  | 20 (7%) | 39 (10%) |
| **Branched PFOS** | |  |  |
|  | Model0 | 1.18 ( 0.62 , 2.27 ) | 0.95 ( 0.55 , 1.64 ) |
|  | Model1 | 1.54 ( 0.77 , 3.14 ) | 0.82 ( 0.44 , 1.54 ) |
| **Linear PFOS** | |  |  |
|  | Model0 | 1.08 ( 0.59 , 1.92 ) | 0.72 ( 0.42 , 1.21 ) |
|  | Model1 | 1.26 ( 0.66 , 2.34 ) | 0.69 ( 0.35 , 1.27 ) |
| **PFHxS** |  |  |  |
|  | Model0 | 1.07 ( 0.65 , 1.63 ) | 0.95 ( 0.62 , 1.38 ) |
|  | Model1 | 1.20 ( 0.70 , 1.93 ) | 0.85 ( 0.50 , 1.34 ) |
| **Octadecanedioic acid** | |  |  |
|  | Model0 | 0.93 ( 0.49 , 1.77 ) | 1.14 ( 0.67 , 1.96 ) |
|  | Model1 | 0.98 ( 0.52 , 1.88 ) | 0.98 ( 0.56 , 1.74 ) |
| **Tridecanedioic acid** | |  |  |
|  | Model0 | 1.09 ( 0.66 , 1.86 ) | 1.34 ( 0.86 , 2.13 ) |
|  | Model1 | 1.11 ( 0.68 , 1.87 ) | 1.37 ( 0.85 , 2.27 ) |
| **Deoxycholic acid** | |  |  |
|  | Model0 | 0.58 ( 0.33 , 1.04 ) | 1.39 ( 0.85 , 2.34 ) |
|  | Model1 | 0.58 ( 0.33 , 1.03 ) | 1.46 ( 0.86 , 2.52 ) |
| ^a^ CI = Confidence Interval; Model0 is the unadjusted model; Model1 is adjusted for maternal age, hospital of delivery, and race/ethnicity; race/ethnicity categories were collapsed due to low numbers of the outcome and to ensure model convergence, categories included: white, Latinx, and other/unknown. | | | |
